# Limited efficacy of 3 mA intensified tDCS of the right inferior frontal cortex for OCD treatment: A randomized, sham-controlled study

**DOI:** 10.1101/2025.04.29.25326694

**Authors:** Atefeh Fatehi-Chenar, Mohsen Dadashi, Alireza Moradi, Soroush Lohrasbi, Abolfazl Ghoreishi, Arash Fazeli, Michael A. Nitsche, Zahra Vaziri, Mohammad Ali Salehinejad

**Author notes:** these authors share last authorship (Zahra Vaziri, Mohammad Ali Salehinejad).

## Abstract

**Objective:** Transcranial direct current stimulation (tDCS) has been proposed for treating obsessive-compulsive disorder (OCD). While it has advantages such as affordability, home-use application, and fewer side effects compared to other treatment options, its clinical efficacy is not yet established, and previous results have been mixed. This randomized, sham-controlled study investigated the efficacy of a novel intensified tDCS protocol targeting the inferior frontal cortex (IFC), a relevant brain region for inhibitory control, for OCD treatment.

**Methods:** Forty patients with OCD received 20 sessions of 3 mA intensified tDCS or sham tDCS in randomized parallel groups. The active treatment included 20 sessions of 20-minute 3 mA stimulation delivered twice per day (in 2 weeks) with 20-min between-session intervals (i.e., intensified). Intervention efficacy and treatment response were evaluated before and after the intervention and at two-week, 1-month, and 3-month follow-ups. Cognitive functions (response inhibition, cognitive flexibility, and sustained attention) were also assessed at these time points. Side effects rating and blinding efficacy were evaluated across groups.

**Results:** Despite higher-rated side effects in the active tDCS group, blinding of patients was successful. OCD symptoms, anxiety, and depression decreased over time in both groups. Despite a significant post vs. pre-intervention reduction of all clinical measures only in the active tDCS group, no statistically significant differences emerged in the active vs sham conditions. The mean change in the OCD symptoms from baseline to the study endpoint and number of responders (≥50% symptom reduction) were 22.38% and 20% (4/20) in the active tDCS and 15.25% and 0.0 (0/20) in the sham tDCS groups. Similarly, cognitive assessments of sustained attention, cognitive flexibility, and response inhibition showed within-group improvements over time but no group-specific effects.

**Conclusions:** The 3 mA intensified tDCS over the right IFC showed limited efficacy confounded with placebo effects for OCD treatment.

## 1. Introduction

Obsessive-compulsive disorder (OCD) is a chronic and debilitating psychiatric condition characterized by persistent, intrusive thoughts (obsessions) and repetitive behaviors or mental acts (compulsions) performed to alleviate associated distress [1]. Affecting approximately 2-3% of the global population, OCD imposes a significant burden on quality of life, impairing social, occupational, and personal functioning [2, 3]. The standard treatments for OCD include cognitive-behavioral therapy (CBT), particularly exposure and response prevention, and pharmacotherapy with selective serotonin reuptake inhibitors (SSRIs). Despite their efficacy, up to 40-60% of patients fail to achieve full symptom remission with these interventions, and many experience intolerable side effects from medications [4, 5]. This treatment gap underscores the need for novel therapeutic strategies for OCD treatment.

Transcranial direct current stimulation (tDCS) has emerged as a promising non-invasive brain stimulation (NIBS) technique with potential applications in psychiatric disorders, including depression, anxiety, and OCD [6–10]. Unlike other forms of neuromodulation, tDCS is portable, low-cost, and safe [11, 12], making it a promising adjunctive treatment for neuropsychiatric disorders [13]. By delivering a weak electrical current through scalp electrodes, tDCS modulates cortical excitability in targeted brain regions, offering a safe and tolerable adjunctive or alternative treatment option [14, 15]. The pathophysiology of OCD involves aberrant activity in cortico-striato-thalamo-cortical (CSTC) circuits, particularly the prefrontal cortex, which regulates inhibitory control and emotional processing [16–18]. Previous studies have explored tDCS as a means to alter and or restore these circuits, with varying degrees of success [19–21].

In recent work [7], we explored the impact of an intensified tDCS protocol at two stimulation intensities (1 and 2 mA) for OCD treatment. The intensified protocol includes 20 minutes of stimulation delivered twice per day with 20-minute between-session intervals, which is able to prolong excitability enhancement [22] and induce superior therapeutic effects in previous studies [8]. In our recent work, we showed that modulation of the prefrontal-supplementary motor network with intensified tDCS ameliorates clinical symptoms of OCD and results in beneficial cognitive effects [7]. That study demonstrated significant reductions in OCD symptoms in the active tDCS groups, but the 2 mA protocol, in particular, showed larger clinical and cognitive effects, indicating that higher intensities within this range may enhance therapeutic outcomes. However, the optimal stimulation parameters—intensity, electrode placement, session frequency, target region—remain under investigation, as subsequent studies have reported inconsistent findings, with some replicating positive effects and others finding no significant difference from sham [19, 21, 23].

Building on this foundation, the current study investigates the efficacy of an intensified tDCS protocol using 3 mA stimulation intensity for OCD treatment. The rationale for increasing stimulation intensity stems from evidence that higher currents may enhance neuromodulatory effects by more robustly altering neuronal excitability and plasticity [24]. In contrast to our previous work, which targeted the right pre-supplementary motor area (pre-SMA) and left dorsolateral prefrontal cortex (DLPFC) with a 1 and 2 mA current [7], this RCT employs a 3 mA current with the anode placed over the right inferior frontal cortex (IFC) and the cathode over the left supraorbital area. This stimulation montage targets the right IFC, a region involved in response inhibition [25, 26]—a core deficit in OCD— and its modulation with anodal stimulation can facilitate response inhibition through dynamic modulation of the fronto-basal ganglia network [27]. Whether and how its modulation, compared to previously targeted regions (e.g., DLPFC, SMA) offers a more direct impact on compulsive behaviors is unknown. Accordingly, in this registered clinical trial (IRCT20230216057437N1), we investigated the impact of intensified tDCS at 3 mA intensity over the right IFC on symptom reduction and cognitive deficits in patients with OCD.

The primary objectives of this study are: (1) to evaluate the efficacy of 3 mA intensified tDCS of the right IFC in reducing OCD symptoms, as measured by the Yale-Brown Obsessive-Compulsive Scale, compared to sham stimulation in a double-blind RCT; (2) to assess its effects on secondary outcomes, including anxiety, depression, and cognitive functions and (3) to evaluate whether and how the right IFC is a promising stimulation target for OCD treatment given the previous mixed results about targeting the pre-supplementary motor area and lateral prefrontal cortex. The novel aspects of this work include (a) the application of 3 mA stimulation in an intensified protocol, (b) and targeting the right IFC for OCD treatment.

## 2. Methods

### 2.1. Participants

This study was a randomized, double-blind, sham-controlled trial with a parallel-group design to prevent blinding failure and carry-over effects. Forty individuals diagnosed with OCD (mean age=35.25, SD=10.56, 23 females) were recruited from several neuropsychiatric clinics in Zanjan, Iran, from April 2023 to August 2024. Patients were randomly assigned to the active (*N*=20) or sham (*N*=20) stimulation groups using the block randomization method (block size of 4). Sample size was calculated a priori based on a medium effect size suggested for tDCS studies (f=0.25, α=0.05, power=0.95, *N*=32, mixed-model ANOVA with 5 measurements). We added 8 more subjects to the total sample size to compensate for potential dropouts, yet none of the participants dropped out before completing the study (Fig. 1). The inclusion criteria were: (1) diagnosis of OCD according to Structured Clinical Interview for DSM-5 (SCID-5) [28], (2) 18-50 years old, (3) non-smoker, (4) no previous history of neurological diseases, brain surgery, epilepsy, seizures, brain damage, head injury, or metal brain implants, and (5) absence of other psychiatric disorders. Those patients on medication from active (*N=6*) and sham (*N*=5) groups were receiving stable doses for at least 6 weeks before the experiment, up to the end of 3^rd^ follow-up. All participants were native speakers and had normal or corrected-to-normal vision. This was a registered clinical trial (IRCT20230216057437N1) approved by the Ethics Committee of the Zanjan University of Medical Science (Ethics code: IR.ZUMS.REC.1401.339). Participants gave their written informed consent before participation (see **Error! Reference source not found.** for demographics).

**Fig. 1:**
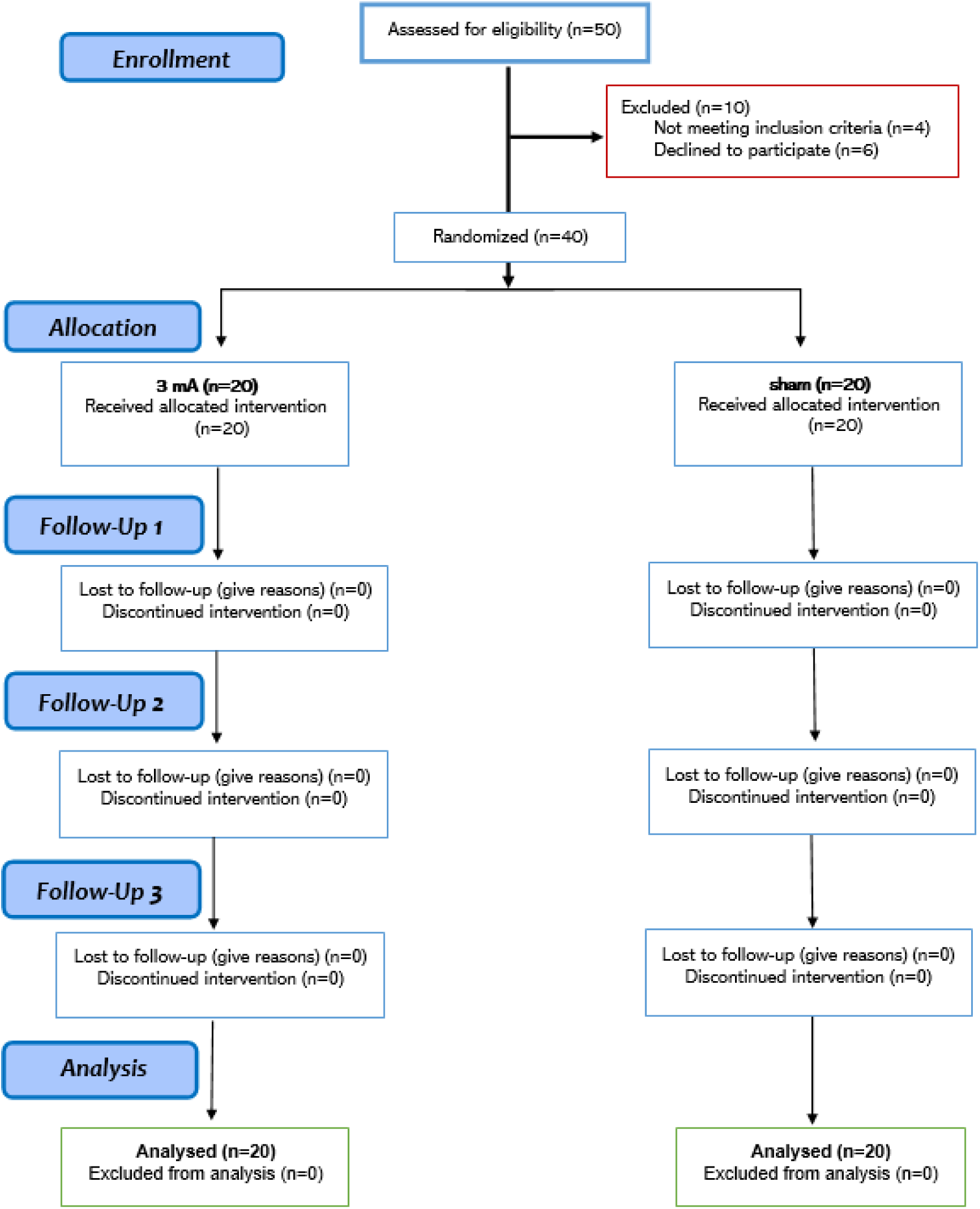
CONSORT flow diagram of study inclusion

### 2.2. Outcome measures

#### 2.2.1. clinical measures

The primary outcome measure to examine the effects of the intervention on OCD symptoms was the Yale-Brown Obsessive-Compulsive Scale (Y-BOCS) [29]. Additionally, anxiety and depressive states were tested by the Beck Anxiety Inventory (BAI) [30] and the Beck Depression Inventory (BDI-II) [31], respectively. Detailed descriptions of measures are in the supplementary content.

#### 2.2.2. cognitive performance

The Cambridge Neuropsychological Test Automated Battery (CANTAB [Cognitive assessment software], 2018) is a computerized collection of cognitive assessments that were used in this study to assess the following areas: cognitive flexibility (assessed through the Intra-Extra Dimensional Set Shift task), response inhibition (evaluated via the Stop Signal Task), and sustained attention (assessed using the Rapid Visual Information Processing task. The CANTAB tests have been thoroughly detailed elsewhere [32, 33]. Detailed descriptions of measures are in the supplementary content.

### 2.1. tDCS

Direct currents were generated by an electrical stimulator (Oasis Pro, MindAlive, Canada) applied through a pair of saline-soaked sponge electrodes (5×5 cm) delivered for 20 minutes on 10 consecutive days (2 sessions per day with 20 min interval, 20 sessions in total). In both active (3-mA) and sham conditions, anodal and cathodal electrodes were placed over the right IFC (F8) and the left supraorbital area (Fp1) respectively, to guarantee a minimum 6 cm distance between the edges of the electrodes [34]. The protocol was adapted from a study demonstrating that anodal tDCS over the right inferior frontal cortex enhances response inhibition by altering functional connectivity within the fronto-basal ganglia inhibitory network [27], which is involved in OCD pathophysiology [26]. In sham stimulation, the electrical current was ramped up and down for 20 seconds each to generate the same sensation as in the active condition and then turned off [35]. A side-effect survey was done after each tDCS session [36]. To guarantee blinding, tDCS was applied by independent investigators who were not involved in outcome measures rating [37]. Blinding efficacy was explored among patients at the end of the last stimulation session by asking participants whether they received a real or a non-real stimulation. After finalizing the study protocol, the patients in the sham group were assigned to active tDCS intervention, but the latter procedure was beyond the focus of the study protocol.

### 2.2. Procedure

Prior to the experiment, participants completed a brief questionnaire to evaluate their suitability for brain stimulation. All participants received 20 sessions of tDCS in 2 weeks. All stimulation sessions took place between 11:00-14:00 and participants were not under sleep pressure [38, 39]. Clinical and cognitive measures were evaluated before the first intervention (pre-intervention), immediately after the end of the last intervention (post-intervention), and also at two-week, four-week, and three-month following the last stimulation session (follow-ups). Patients were instructed about the tasks before the beginning of the experiment and the task stimuli order was randomized in each measurement. None of the patients received any kind of psychotherapy during the study. Participants were blind to the study hypotheses and stimulation conditions. The experimenter who conducted the outcome measures was blinded to the tDCS conditions (Fig. 2).

**Fig. 2:**
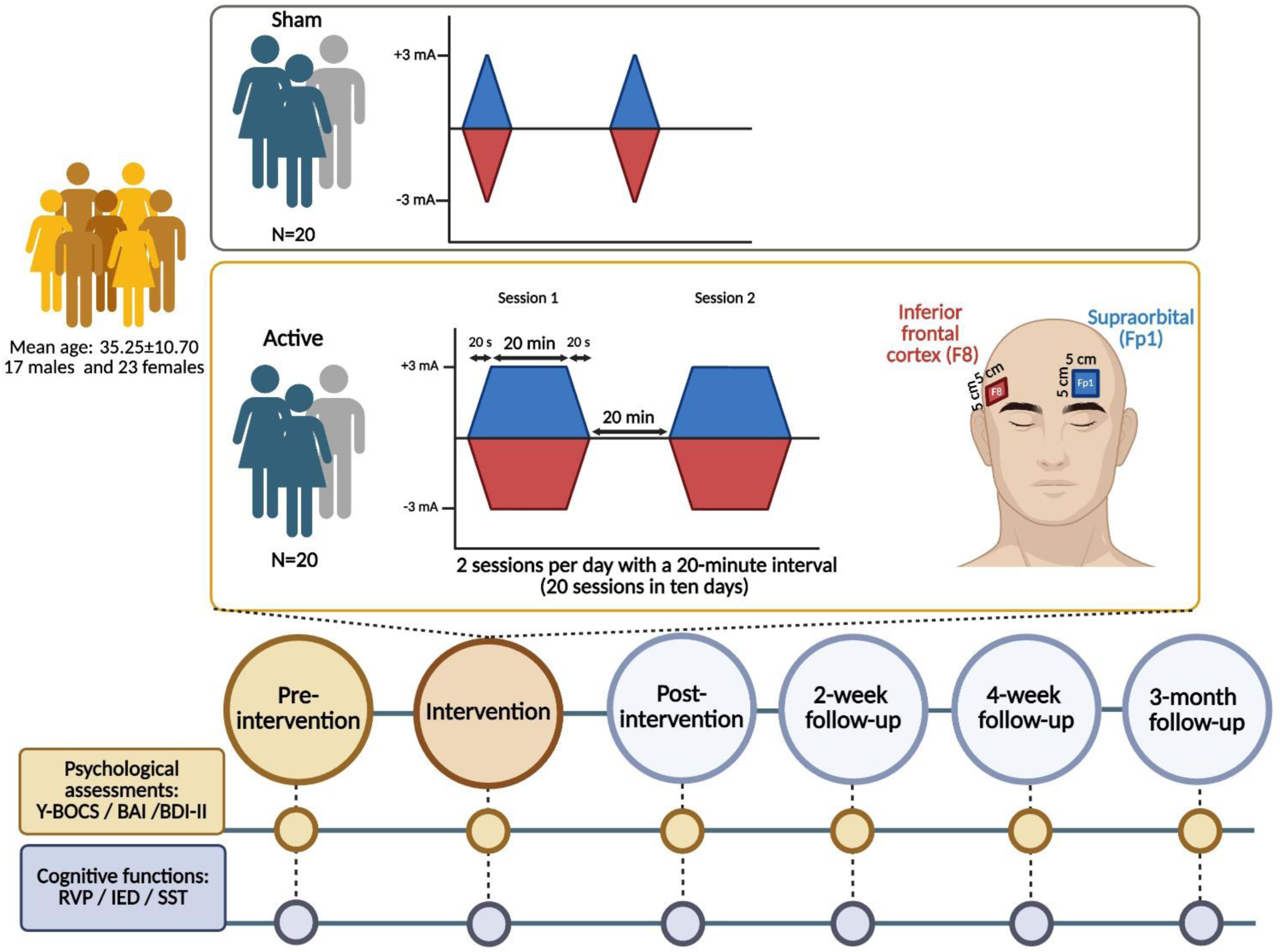
Experimental procedure

### 2.3. Statistical analysis

Data was analyzed with the statistical package SPSS, version 27.0 (IBM, SPSS, Inc., Chicago, IL) and GraphPad Prism 9.1 (GraphPad Software, San Diego, California). The normality and homogeneity of data variance were confirmed by Shapiro-Wilk and Levin tests, respectively. Mixed model ANOVAs were conducted for the dependent variables (clinical measures: obsessive-compulsive, depression, and anxiety symptoms; cognitive measures: sustained attention, cognitive flexibility, response inhibition) with “group” (active vs sham) as the between-subject factor and time (pre-intervention, post-intervention, two-week, four-week, and three-month follow-ups) as the within-subject factor. Mauchly’s test was used to evaluate the sphericity of the data before performing the respective ANOVAs (*p*<0.05). In case of violation, degrees of freedom were corrected using the Greenhouse-Geisser method. In the event of significant findings in the ANOVAs, LSD post hoc comparisons were executed across time points for each group and between-group for each time point. The association between OCD subtypes and symptom reduction was investigated the Pearson’s correlational analysis. Blinding efficacy was examined with the chi-squared test for Independence on participants’ guesses of blinding (0, 1) and Bang’s Blinding Index (−1, 1) [40], with negative values suggesting that participants frequently guessed the opposite of their actual treatment. The critical level of significance was 0.05 for all statistical analyses.

## 3. Results

### 3.1. Baseline assessment, safety outcomes, and blinding efficacy

**Error! Reference source not found.** summarizes demographic data, with no significant differences observed across groups. **Error! Reference source not found.** and S2 show the mean and standard deviation of outcome measures (overall and symptom category), with no significant between-group differences in baseline measurements of outcome variables. One-way ANOVAs were conducted on average reported side effects across groups. The results revealed significantly higher ratings of itching (*F*_1,38_= 20.10, *p*<0.001), burning (*F*_1,38_= 47.53, *p*<0.001), pain (*F*_1,38_= 12.36, p=0.001), and skin redness (*F*_1,38_= 28.21, *p*<0.001) in the active tDCS group vs the sham group. No significant difference was found in trouble concentrating between the two groups (*F*_(1,38)_= 0.114, *p*=0.738) (Table S3). Regarding blinding efficacy, 3 out of 20 participants (15%) in the active group noticed their condition correctly, while 17 out of 20 (85%) did not. In the sham group, 4 out of 20 participants (20%) noticed their condition correctly, whereas 16 out of 20 (80%) did not. The results of the Chi-Square test (χ2=0.173; p=0.677) and Bang’s Blinding Index (active = −0.7; sham= −0.6) indicated that the blinding was successful, suggesting that participants’ awareness of their tDCS condition did not significantly influence their perception of side effects.

### 3.2. Clinical Outcome Measures

#### 3.2.1. OCD symptoms

For Y-BOCS scores, a significant main effect of time (*F*_3.276,124.470_*=*13.884, *p*<.001, *ηp²=*0.268) but no main effect of group (*F*_1,38_*=*0.234, *p*=0.0632, *ηp²=*0.006) or the time×group interaction (*F*_3.276,124.470_*=*0.326, *p*=0.823, *ηp²=*0.009) were observed. Within-group post-hoc LSD comparisons indicated significant Y-BOCS score reductions from baseline in the active-tDCS group at post-intervention (*p*=0.023), 2-week (*p*=0.015), and 3-month follow-ups (*p*=0.023). The sham-tDCS group showed significant reductions only at the 2-week (*p*=0.031) and 3-month follow-ups (*p*=0.033). No significant between-group differences were found at any time point, including the baseline differences.

#### 3.2.2. Anxiety state

For the BAI scores similarly, a significant main effect of time (*F*_4,152_ *=* 13.221, *p*<.001, *ηp²=* 0.258) and no effect of group (*F_1,38_*=1.542, *p*=0.222, *η*p^2^=0.039) and time×group interaction (*F_4,152_*=0.728, *p*=0.574, *η*p^2^=0.019) were observed. Post-hoc LSD comparisons indicated a significant decrease in BAI scores only in the active-tDCS group at post-intervention *(p*=0.018), 2-week *(p*=0.020), 4-week *(p*=0.023), and at 3-month (*p*=0.016) follow-ups vs pre-intervention. Regarding between-group analysis, no significant differences were observed at any time point.

#### 3.2.3. Depressive state

BDI scores showed a significant main effect of time (*F*_4,152_ *=* 6.635, *p*<.001, *ηp²=* 0.149) and no main effect of group (*F_1,38_*=1.854, *p*=0.181, *η*p^2^=0.047) or time×group interaction (*F_4,152_*=0.734, *p*=0.570, *η*p^2^=0.019). Within-group post hoc LSD comparisons showed a significant reduction in BDI scores only in the active-tDCS group at post-intervention *(p*=0.036) compared to the pre-intervention. Between-group analysis revealed no significant differences between the active and sham tDCS groups at any time point, including the baseline differences.

#### 3.2.4. Symptom reduction and treatment response

We also investigated the average symptom reduction treatment response (defined as at least 50% symptom reduction after the intervention) across groups. For OCD symptoms, the active tDCS group showed an average reduction of 22.38%, with 20% of patients responding, compared to a 15.25% symptom reduction and 0% response in the sham group. For BAI scores, active tDCS led to a 32.58% average reduction and a 40% response rate, while the sham group showed a reduction of 26.73% and a 20% response rate, suggesting a partial placebo effect. For depressive symptoms, active tDCS and sham groups had average reductions of 19.33% and 12.42%, respectively, with response rates of 20% and 15%, respectively, suggesting a partial placebo effect. Our correlational analyses of treatment response and OCD subtypes did not show any significant association between symptom-specific treatment response.

### 3.3. Cognitive outcome measures

#### 3.3.1. The Rapid Visual Information Processing (RVP)

Sustained attention was assessed with the RVP with RVP-A (accuracy of target detection adjusted for false positives), hits (accuracy), and latency of correct responses as major outcome measures. The mixed-model ANOVA revealed a significant main effect of time on RVP-A scores (*F*_4,152_*=* 9.462, *p<* 0.001, *ηp²=*0.199), but no significant main effect of group (*F*_1,38_*=* 0.148, *p*=0.702, *ηp*²=0.004) or time×group interaction (*F*_4,152_*=* 1.025, *p*=0.396, *ηp*²=0.026). Post hoc analyses showed significantly higher RVP-A scores in the active tDCS group at the 2-week (*p*=0.020) and 4-week (*p*=0.046) follow-ups compared to pre-intervention, and in the sham tDCS group at the 3-month follow-up (*p*=0.032). Analysis of accuracy (hits) similarly showed a significant main effect of time (*F*_4,152_ *=* 9.160, *p*<0.001, *ηp²=*0.194) with non-significant group (*F*_1,38_ *=* 0.004, *p*=0.952, *ηp²=*0.000) and interaction effects (*F*_4,152_ *=* 1.010, *p*=0.404, *ηp²=*0.026). Hits significantly increased in the active-tDCS group at the 2-week follow-up (*p*=0.016) and in the sham tDCS group at the 3-month follow-up (*p*=0.015) relative to pre-intervention. For latency, there was also a significant main effect of time (*F*_4,120_ *=* 6.925, *p*<0.001, *ηp²=*0.188), while group (*F*_1,30_ *=* 0.205, *p*=0.654, *ηp²=*0.007) and interaction effects (*F*_4,120_ *=* 0.715, *p*=0.583, *ηp²=*0.023) were not significant. Post hoc tests revealed significantly reduced latency in the active tDCS group at post-intervention (*p*=0.041), 2-week (*p*=0.036), 4-week (*p*=0.032), and 3-month follow-ups (*p*=0.001) compared to pre-intervention. No significant latency changes were observed in the sham tDCS group. No significant between-group differences were found for RVP-A, hits, or latency.

#### 3.3.2. Intra-Extra Dimensional Set Shift (IED)

Cognitive flexibility was measured with the IED. The ANOVA revealed a significant main effect of time for latency (*F*_3.032,115.214_*=* 16.531, *p*<0.001, *ηp²=*0.303) and stages completed (*F*_4,152_ *=* 3.366, *p*=0.011, *ηp²=*0.081). Post hoc analyses revealed significant latency reductions from baseline in both the active-tDCS group (2-week: *p*=0.0004; 4-week: *p*=0.0001; 3-month: *p*=0.0004) and the sham-tDCS group (2-week: *p*=0.008; 4-week: *p*=0.008; 3-month: *p*=0.030). Significant effects for stages-completed did not survive pairwise comparison. The effect of group was not significant for latency (*F* _(1,38)_ *=* 0.633, *p*=0.431, *ηp²=* 0.016) and stages completed (*F* _(1,38)_ *=* 2.132, *p*=0.152, *ηp²=* 0.053). The time×group interaction also showed no significant effects on total latency (*F_3.03,115.21_*=1.268, *p*=0.289, *η*p^2^=0.032), and stages-completed (*F_4,152_*=0.435, *p*=0.783, *η*p^2^=0.011).

#### 3.3.3. Stop Signal Task (SST)

Response inhibition was measured by the SST. The ANOVA revealed no significant main effects of time or group, nor a significant time x group interaction, for any outcome measure (accuracy, reaction time, or proportion of successful stops). The detailed statistical values are as follows: Time – accuracy (*F_2.70,102.90_*=0.399, *p*=0.733, *η*p^2^=0.010), reaction time (*F_2.60,98.82_*=1.362, *p*=0.261, *η*p^2^=0.035), and proportion of successful stops (*F_4,152_*=0.447, *p*=0.774, *η*p^2^=0.012). Main effect of group: accuracy (*F_1,38_*=1.473, *p*=0.232, *η*p^2^=0.037), reaction time (*F_1,38_*=0.394, *p*=0.534, *η*p^2^=0.010), and proportion of successful stops (*F_1,38_*=0.563, *p*=0.458, *η*p^2^=0.015). time×group interaction: accuracy (*F_2.708,102.904_*=0.103, *p*=0.947, *η*p^2^=0.003), reaction time (*F_2.601,98.820_*=0.390, *p*=0.732, *η*p^2^=0.010), and proportion of successful stops (*F_4,152_*=0.143, *p*=0.966, *η*p^2^=0.004).

## 2. Discussion

This randomized, double-blind, sham-controlled trial investigated the efficacy of 3 mA intensified tDCS over the right IFC for treating OCD in 40 patients. The primary finding was a significant reduction in OCD symptoms (Y-BOCS scores), anxiety (BAI scores), and depression (BDI-II scores) over time in both the active and sham tDCS groups. Despite a significant post vs. pre-intervention reduction of all clinical measures only in the active tDCS group, and a higher number of responders in the active group (Fig. 3), no statistically significant differences emerged between the groups (active vs sham) across clinical outcome measures. Similarly, cognitive assessments of sustained attention, cognitive flexibility, and response inhibition showed within-group improvements over time but no group-specific effects. These results suggest that the intensified 3 mA anodal tDCS right IFC-cathodal left supraorbital did not provide a therapeutic advantage beyond placebo effects. In what follows, we first discuss the results with respect to target region and stimulation intensity, two novel aspects of the current study. We then discuss the observed placebo effects and potential reasons and finally compare the current 3 mA intensified tDCS protocol with the current tDCS literature in OCD.

**Fig. 3:**
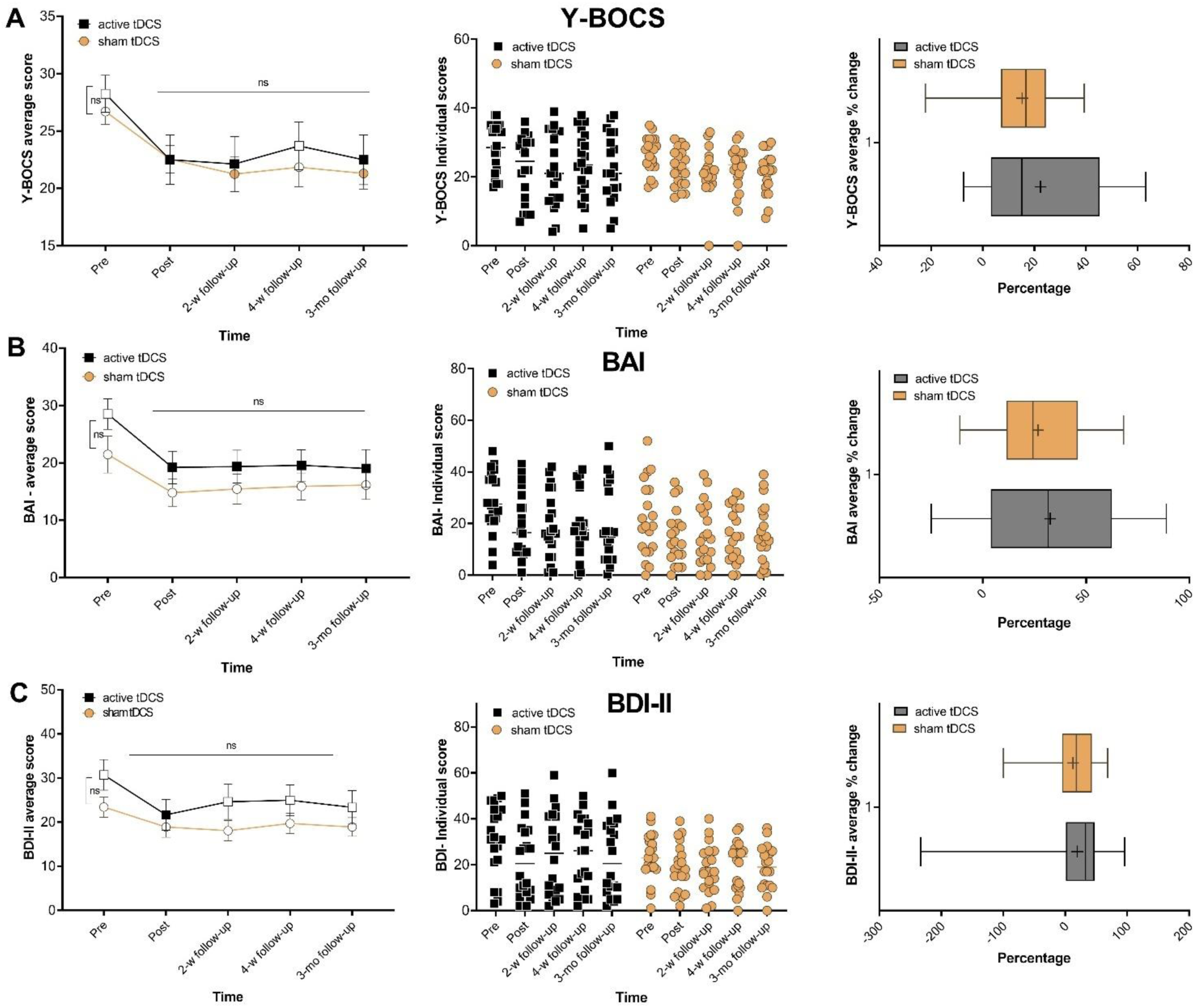
OCD symptoms, depression and anxiety states were measured by the Y-BOCS (A), BAI (B), and BDI-II (C) before, after, and up to two weeks, 1 month, and 3 months following the intervention. In the left panel, filled symbols indicate significant differences at each time point compared to pre-intervention scores. The middle panel displays a scatter plot of clinical scores for each group across time points. The right panel shows the mean score change from the baseline to the study endpoint (week 2) for the respective measure. The horizontal bar shows the median, the + shows the mean, the upper and lower boundaries show the 25th and 75th percentiles, respectively, and the whiskers show the 1– 99th percentile. Pairwise comparisons were conducted with post-hoc LSD tests, and error bars indicate the standard error of the mean (s.e.m.). *Note*: Y-BOCS = Yale-Brown Obsessive-Compulsive Scale; BAI = Beck Anxiety Inventory; BDI-II = Beck Depression Inventory-II; tDCS = transcranial direct current stimulation; w = week; mo = month; ns = non-significant.

**Fig. 4:**
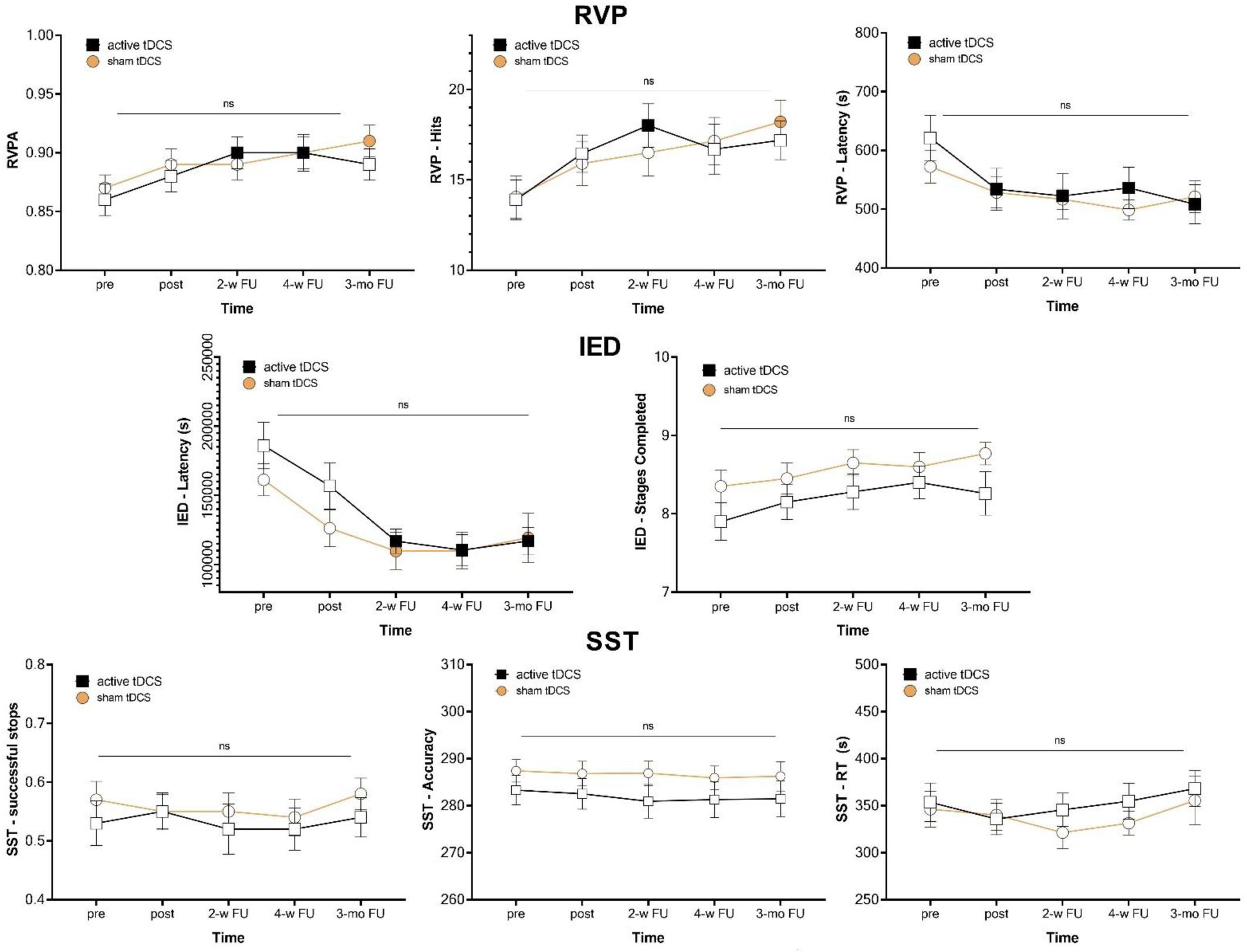
Cognitive performance was evaluated before and after the 10^th^ session of 3 mA intensified and sham tDCS using the CANTAB neuropsychological test battery for OCD, which includes measures of sustained attention (RVP), cognitive flexibility (IED), and response inhibition (SST). Filled symbols indicate significant differences at each time point compared to pre-intervention scores. Post hoc tests were conducted with LSD pairwise comparisons, and error bars indicate the standard error of the mean (s.e.m.). *Note*: tDCS = transcranial direct current stimulation; RVP = Rapid Visual Processing; IED = Intra-Extra Dimensional Set Shift; SST = Stop Signal Task; s = second; w = week; mo = month; FU = follow-up; ns = non-significant.

### 2.1. Limited efficacy: Relevance of target region and stimulation intensity

#### 2.1.1. Is the right IFC a reliable target region for OCD treatment?

One novel aspect of our tDCS protocol was targeting the right IFC with anodal tDCS based on the previous studies in healthy humans that have shown its involvement in response inhibition [25] and specifically the study that showed anodal tDCS over the right IFC facilitates response inhibition and functional connectivity between the right pre-SMA and subthalamic nuclei [27], two highly-involved regions in OCD pathophysiology [26, 41]. Anodal stimulation of the right IFC and the cathodal stimulation of the left supraorbital area were expected to target inhibitory control circuits involved in OCD. The orbitofrontal cortex (OFC) is anatomically very close the the right IFC, and is hyperactive in OCD patients [41]. It is possible that the 3 mA anodal stimulation, with its diffuse induced electrical field, has also increased excitability in the OFC or failed to sufficiently regulate hyperactivity in the cortico-striato-thalamo-cortical circuit, which contributes to OCD thoughts and behavior [41]. The modulation of the left frontopolar with cathodal stimulation should not be ignored. Although inhibiting the left OFC was anticipated, cathodal tDCS at higher intensities may not achieve this due to the complexities involved [42]. This might partially explain the limited therapeutic efficacy of the intervention.

Furthermore, none of the outcome measures of SST (response inhibition task) improved in both groups, indicating that the intervention failed to enhance inhibitory control in OCD patients. Although 20% of the active group (4/20) achieved a ≥50% Y-BOCS reduction, compared to 0% in the sham group, the mean symptom reductions (22.38% active vs. 15.25% sham) and lack of statistical significance of both symptoms and response inhibition performance suggest that this configuration or the 3 mA intensity may not have optimally engaged the relevant neural networks.

#### 2.1.2. 3 mA tDCS could be too much

The limited efficacy of 3 mA tDCS may also stem from neurophysiological effects of higher stimulation intensities. In our study, 3 mA tDCS over the left frontopolar cortex and right IFC may have induced counterproductive effects. Importantly, in a tDCS study conducted on motor cortical excitability, the same intervention (3 mA anodal tDCS with 20 min interval) was conducted on motor cortical excitability and the author found that in contrast to 1 mA tDCS that prolonged after effects for 24 h, the 3 mA tDCS after effects were limited to 120 minutes [43]. This study, most relevantly explains the limited efficacy of our intervention not just failed upregulation of the right IFC, but also failed downregulation of the left frontopolar cortex (Fp1), a nearby region to OFC.

A hypothesized mechanism here is calcium overflow as a result of 3 mA stimulation. While moderate increases in intracellular calcium facilitate long-term potentiation (LTP), excessive calcium influx can trigger depotentiation or long-term depression (LTD), preventing further plasticity induction [44, 45]. It is possible that intensified tDCS of the prefrontal regions, as shown in the motor cortex [43], has a stronger activating effect on NMDAR, and the resulting higher amount of calcium influx induces counterbalancing effects with a second stimulation [46]. Furthermore, we applied the 3 mA tDCS in intensified mode (20 min stimulation + 20 min rest + 20 min stimulation), which was shown to limit later LTP compared to 1 mA [43]. The 2^nd^ stimulation period was during the aftereffects of the first period, which could prolong excitability in previous tDCS studies in motor excitability only in 1 and mA but not 3 mA intensity [22, 43, 47]. While in our previous work, we showed a significantly larger therapeutic efficacy of the 2 mA intensified tDCS vs 1 mA [7], the 3 mA intensity might have been excessive. This comparison is, however, limited by the different target regions used in our previous study (left DLPFC and right pre-SMA).

### 2.2. The placebo effect

The observed improvements in both active and sham groups indicate a placebo effect, consistent with prior NIBS studies. A meta-analysis of rTMS trials in depression found a large placebo response linked to depression improvement [48]. In tDCS studies, similar antidepressant effects were reported [49]. In treatment-resistant OCD, two recent studies found that active tDCS was not superior to sham despite symptom reduction [19, 50]. In one study, 20 daily sessions of cathodal SMA tDCS significantly reduced OCD symptoms compared to sham, but no between-group differences were found in treatment response and anxiety/depression symptom reduction [50]. The other study with 10 sessions of 2 mA tDCS (cathodal SMA-anodal Fp2) reported a significant Y-BOCS score decrease over time in both groups [19]. These findings parallel our study, where the sham group’s 15.27% reduction in Y-BOCS scores and 26.73% reduction in BAI scores suggest the presence of a placebo response.

The placebo effect is influenced by multiple factors, including patients’ belief in the treatment, trust in healthcare providers, ritualistic treatment procedures, and the therapeutic context, particularly for those with prior unsuccessful treatments. Patients’ expectations of improvement may alter neurophysiology and emotional states [51], which may contribute to OCD symptom reduction. Clinician-rated scales show a stronger placebo effect than self-reported measures [52]. Furthermore, NIBS techniques, like tDCS and rTMS, may produce greater placebo effects compared to other treatments [53]. Lastly, a meta-analysis of OCD RCTs (regardless of intervention type) showed that placebos are about 50% as effective as active interventions in OCD trials, especially in younger patients [52]. Our results also highlight these factors, indicating that placebo effects may have been underestimated in prior tDCS studies for OCD.

### 2.3. Comparison with existing tDCS studies in OCD

Previous tDCS studies in OCD have mostly used 1-2 mA stimulation intensity over other regions (e.g., dorsolateral prefrontal cortex, pre-SMA), and results have been inconsistent. Some studies reported modest symptom reductions, mostly with cathodal pre-SMA and anodal left DLPFC tDCS [7, 21, 23], while others (targeting the pre-SMA) found no difference from sham [19, 50]. The current study’s use of 3 mA extends this exploration but suggests that this specific tDCS configuration (3 mA anodal right IFC-cathodal Fp1) showed limited clinical efficacy for reducing OCD symptoms and improving response inhibition. The excessive heterogeneity in OCS pathophysiology [41, 54], clinical predictors of response [55], the impact of stimulation parameters (including intensity and target region) and OCD subtypes [56] are contributing factors to NIBS treatment response in OCD that could affect the limited efficacy of our intervention although OCD subtypes did not correlate with treatment response in our sample.

### 2.4. Limitations, strengths, and future Directions

Limitations include the absence of neuroimaging and/or neurophysiological data that could have provided more information about the neurophysiological effects of the intervention across groups. Second, our study was monocentric, limiting its generalizability. The strengths of our study were careful monitoring of symptom and cognitive performance changes across 5 time-points, and assessment of cognitive functions in addition to clinical assessment. Future larger, multi-site trials with titration of stimulation parameters (intensity, polarity) along with neuroimaging data could clarify the mechanisms of effects for right IFC tDCS for OCD treatment.

### 2.5. Conclusion

To conclude, this sham-controlled RCT found no evidence that 3 mA intensified tDCS over the right IFC outperforms sham stimulation for OCD treatment. Strong placebo effects, as seen in other tDCS studies, insufficient or ineffective induced excitability alteration in the target region, or suboptimal stimulation parameters likely contributed to the lack of differential outcomes across groups. Further tDCS studies for therapeutic use in OCD with titration of parameters of space are needed, especially by focusing on more promising target regions.

**Table 1.**
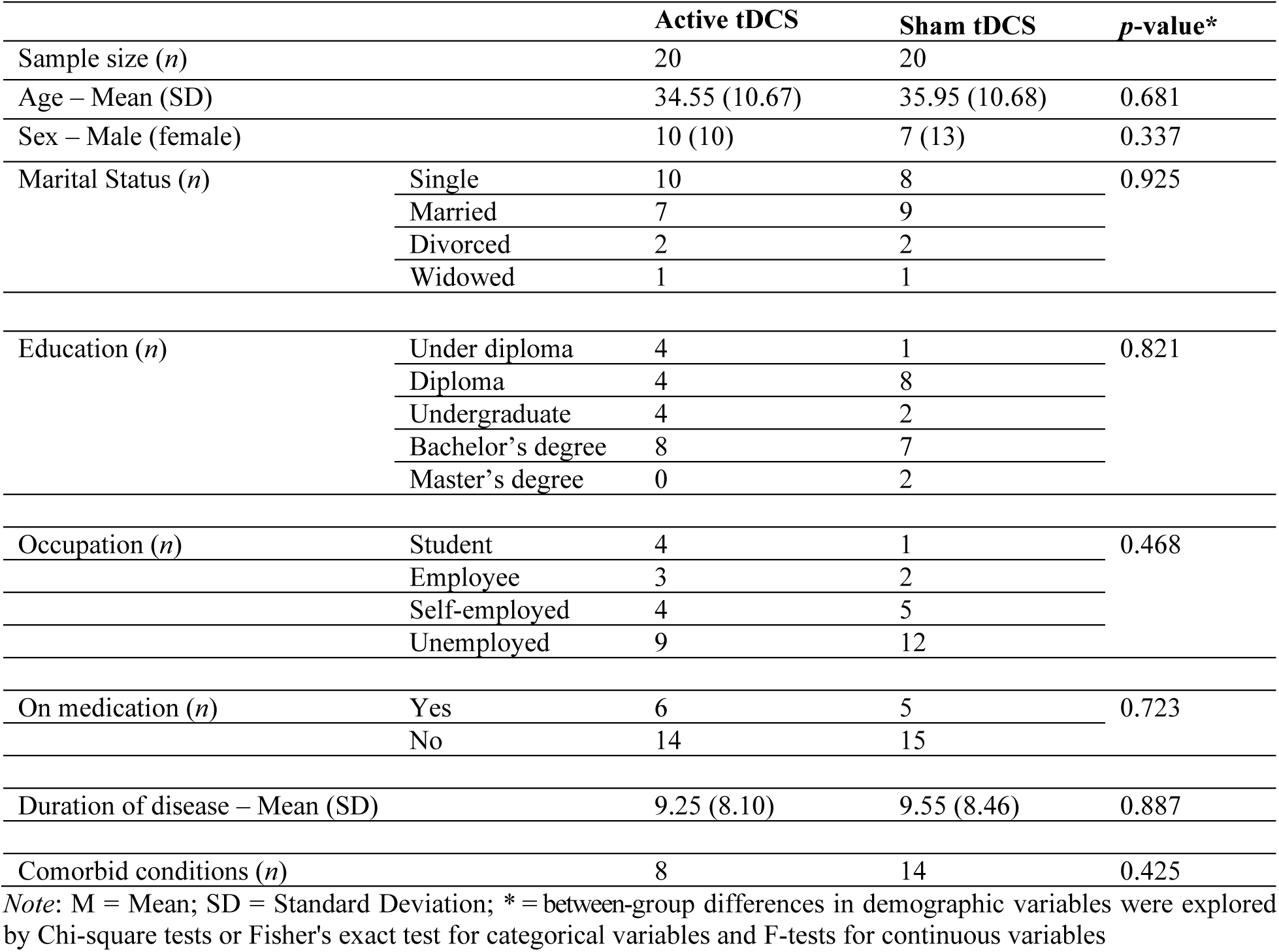
Demographic data.

|  |  | **Active tDCS** | **Sham tDCS** | ***p*-value\*** |
| --- | --- | --- | --- | --- |
| Sample size (*n*) |  | 20 | 20 |  |
| Age – Mean (SD) |  | 34.55 (10.67) | 35.95 (10.68) | 0.681 |
| Sex – Male (female) |  | 10 (10) | 7 (13) | 0.337 |
| Marital Status (*n*) | Single | 10 | 8 | 0.925 |
|  | Married | 7 | 9 |  |
|  | Divorced | 2 | 2 |  |
|  | Widowed | 1 | 1 |  |
| Education (*n*) | Under diploma | 4 | 1 | 0.821 |
|  | Diploma | 4 | 8 |  |
|  | Undergraduate | 4 | 2 |  |
|  | Bachelor's degree | 8 | 7 |  |
|  | Master's degree | 0 | 2 |  |
| Occupation (*n*) | Student | 4 | 1 | 0.468 |
|  | Employee | 3 | 2 |  |
|  | Self-employed | 4 | 5 |  |
|  | Unemployed | 9 | 12 |  |
| On medication (*n*) | Yes | 6 | 5 | 0.723 |
|  | No | 14 | 15 |  |
| Duration of disease – Mean (SD) |  | 9.25 (8.10) | 9.55 (8.46) | 0.887 |
| Comorbid conditions (*n*) |  | 8 | 14 | 0.425 |

*Note:* M = Mean; SD = Standard Deviation; \* = between-group differences in demographic variables were explored by Chi-square tests or Fisher's exact test for categorical variables and F-tests for continuous variables

## Data Availability

All data produced in the present study are available upon reasonable request to the authors

## Acknowledgments

MAN is supported by the Deutsche Forschungsgemeinschaft (DFG, German Research Foundation)—Project Number 316803389—SFB 1280, project A6, and by the German Centre of Mental Health (Project Number 01EE2302D). We would like to thank Dr. Ali Fathi Jouzdani for his technical support.

## Credit Author Contributions

**A-F**: Investigation, Formal analysis (support), Data curation, Validation. **MD**: Supervision, Resources, Project administration, Validation. **AM & SL**: Software (Cognitive assessment). **A-GH & AF**: Project administration (patient diagnosis and allocation). **Michael A. Nitsche:** Supervision, Writing - review & editing. **ZV**: Formal analysis (lead), Visualization, Writing - Original draft (support). **MAS**: Conceptualization, Methodology, Supervision, Visualization, Writing - Original draft (lead), Writing - Review and Editing.

